# Evaluation of the diagnostic value of YiDiXie™-SS, YiDiXie™-HS and YiDiXie™-D in lung cancer

**DOI:** 10.1101/2024.07.18.24310612

**Authors:** Shengjie Lin, Xutai Li, Wuping Wang, Chen Sun, Zhenjian Ge, Wenkang Chen, Yingqi Li, Yutong Wu, Pengwu Zhang, Siwei Chen, Huimei Zhou, Wei Li, Hua Chen, Jixian Liu, Yongqing Lai

## Abstract

**Background:** Lung cancer poses a serious threat to human health. Lung CT is widely used for lung tumor screening or initial diagnosis, while contrast-lung CT is extensively employed in lung tumor diagnosis. However, false-positive lung CT results can lead to misdiagnosis and incorrect surgery or treatment, while false-negative lung CT results can lead to missed diagnosis and delayed treatment. There is an urgent need to find convenient, cost-effective and non-invasive diagnostic methods to reduce the false-positive rate and false-positive rates of lung CT. The aim of this study was to evaluate the diagnostic value of YiDiXie™-SS, YiDiXie™-HS and YiDiXie™-D, in lung cancer.

**Patients and methods:** This study finally included 1250 subjects (the malignant group, n=1078; the benign group, n=172). Remaining serum samples from the subjects were collected and tested using the YiDiXie™ all-cancer detection kit, which was applied to assess the sensitivity and specificity of YiDiXie™-SS, YiDiXie™-HS and YiDiXie™-D, respectively. **Results**: YiDiXie™-SS has a sensitivity of 97.5% (96.4% - 98.3%) and its specificity is 66.9% (59.5% - 73.5%). This means that YiDiXie™-SS has very high sensitivity and high specificity in lung tumors.YiDiXie™-HS has a sensitivity of 85.3% (83.0% - 87.2%) and its specificity is 83.1% (76.8% - 88.0%). This means that YiDiXie™-HS has high sensitivity and high specificity in lung tumors.YiDiXie™-D has a sensitivity of 73.9% (71.2% - 76.5%) and its specificity is 92.4% (87.5% - 95.5%). This means that YiDiXie™-D has high sensitivity and very high specificity in lung tumors.YiDiXie™-SS has a sensitivity of 98.2% (95% CI: 96.8% - 99.0%) and a specificity of 66.0% (95% CI: 51.7% - 77.8%) in patients with CT-positive tumors. This means that the application of YiDiXie™-SS reduces the CT false-positive rate by 66.0% (95% CI: 51.7% - 77.8%) with essentially no increase in malignancy underdiagnosis. The sensitivity of YiDiXie™-HS in CT negative patients was 84.2% (95% CI: 80.6% - 87.2%) and its specificity was 82.4% (95% CI: 74.8% - 88.1%). This means that the application of YiDiXie ™ -HS reduces the false-negative lung CT rate by 84.2% (95% CI: 80.6% - 87.2%). YiDiXie™-D has a sensitivity of 73.7% (70.1% - 77.1%) and a specificity of 93.6% (82.8% - 97.8%) in CT-positive patients. This means that YiDiXie ™ -D reduces the rate of CT false positives by 93.6% (82.8% - 97.8%). YiDiXie™-D has a sensitivity of 74.2% (70.1% - 78.0%) and a specificity of 92.0% (85.9% - 95.6%) in patients with a negative CT. This means that YiDiXie™-D reduces the CT false-negative rate by 74.2% (70.1% - 78.0%) while maintaining high specificity.

**Conclusion:** YiDiXie™-SS provides extremely high sensitivity and high specificity in lung tumors. YiDiXie™-HS provides high sensitivity and high specificity in lung tumors. yiDiXie™-D provides high sensitivity and very high specificity in lung tumors. YiDiXie ™ -SS significantly reduces the rate of lung CT false positives with essentially no increase in delayed treatment of malignant tumors. YiDiXie™-HS significantly reduces the false-negative rate of lung CT. YiDiXie™-D can significantly reduce the false-positive rate of lung CT or significantly reduce the false-negative rate while maintaining high specificity. the YiDiXie™ test has important diagnostic value in lung cancer and is expected to solve the problems of “too high false-positive rate” and “too high false-negative rate” of lung CT.

**Clinical trial number:** ChiCTR2200066840.

## INTRODUCTION

Lung cancer is one of the most common malignant tumors^1,2^. According to the latest data, the number of new lung cancer cases reached 2.5 million and the number of new deaths was 1.8 million in 2022 globally^2^. Compared with 2020, lung cancer incidence and mortality in 2022 increased by 12.4% and 1.2%, respectively^1,2^. Due to the lack of typical clinical symptoms, the majority of lung cancer patients are diagnosed at an advanced stage with extensive metastasis of the tumor, thus missing out on optimal treatment. The five-year survival rate of lung cancer in most countries around the world is 10-20%^3^, while the five-year survival rate of distant metastasis in small cell lung cancer is less than 5%^4^. Thus, lung cancer is a serious threat to human health.

Lung CT is widely used in the screening or diagnosis of lung cancer. On the one hand, lung CT can produce a large number of false-positive results. According to the National Lung Screening Trial Study (NLST), the false-positive rate was 23.3% when defining a nodule >4 mm as a positive screening^5^. A Chinese study also confirmed that 804 participants screened positive under the NLST criteria, but only 51 were diagnosed with lung cancer at the two-year follow-up, a false-positive rate as high as 21.8%^6^. A study of 246 positive screening non-small cell lung cancer cases found that 141 were false positives, with a positive predictive value of only 43% and a false-positive rate of 29%^7^. The false positive rate increased to 41.5% when the scan lasted longer than 4 minutes^8^. In a study focusing on the characterization of lung nodules by lung CT, the false positive rate was 71%^9^. With a positive lung CT, patients usually undergo tumor resection or radical resection^10^. Positive enhancement CT results may lead to benign diseases being misdiagnosed as malignant tumors, and patients may face undesirable consequences such as unnecessary emotional distress, costly surgeries and investigations, surgical trauma, and even organ removal and loss of function. Therefore, there is an urgent need to find a convenient, cost-effective, and noninvasive diagnostic method to reduce the false-positive rate of lung CT.

On the other hand, lung CT can also produce a large number of false-negative results. In a study of non-small cell lung cancer, the false negative rate of lung CT was found to be 31%^7^. A multicenter study showed an 18.9% false-negative rate on lung CT for malignant lung nodules when patients had intravenous contrast injected for >1 minute^8^. When enhancement CT is negative, patients are usually taken for observation and regular follow-up^10^. Negative enhancement CT results may lead to malignant tumors being misdiagnosed as benign diseases, delaying the timing of treatment and progression of the disease to an advanced stage, and as a result, patients may face adverse consequences such as poor prognosis, high cost of treatment, decreased quality of life, and shorter survival. Therefore, there is an urgent need to find a convenient, cost-effective and noninvasive diagnostic method to reduce the false-negative rate of lung CT.

In addition, there are some special patients who need to be extra cautious in choosing whether or not to operate, such as: smaller tumors, tumors that require lobectomy, patients with significant pulmonary insufficiency, and patients in poor general condition. The risk of wrong surgery in these special patients is much higher than the risk of missed diagnosis. And false-positive CT results mean that benign diseases are misdiagnosed as malignant tumors, which will lead to misdiagnosis and wrong surgery. Therefore, there is an urgent need to find a convenient, cost-effective and noninvasive diagnostic method with high specificity to substantially reduce the false-positive rate of lung CT in these special patients or to significantly reduce its false-negative rate while maintaining high specificity.

Based on the detection of miRNAs in serum, Shenzhen KeRuiDa Health Technology Co., Ltd. has developed “YiDiXie ™ all-cancer test” (hereinafter referred to as the YiDiXie ™ test)^11^. With only 200 milliliters of whole blood or 100 milliliters of serum, the test can detect multiple cancer types, enabling detection of cancer at home^11^. The YiDiXie ™ test consists of three independent tests: YiDiXie ™ -HS, YiDiXie™-SS and YiDiXie™-D^11^.

The purpose of this study is to evaluate the diagnostic value of YiDiXie™-SS, YiDiXie™-HS and YiDiXie™-D in lung tumors.

## PATIENTS AND METHODS

### Study design

This work is part of the sub-study “Evaluating the diagnostic value of the YiDiXie ™ test in multiple tumors” of the SZ-PILOT study (ChiCTR2200066840).

SZ-PILOT is a single-center, prospective, observational study (ChiCTR2200066840). Subjects who gave informed consent for the donation of their residual samples at the time of admission or during physical examination were taken into consideration for inclusion. For this study, a serum sample of 0.5 milliliters was obtained.

Participants in this study were kept blinded. Laboratory professionals conducting the YiDiXie™ Test and KeRuiDa laboratory technicians analyzing the test results were unaware of the clinical information of the participants. Clinical specialists evaluating the individuals’ clinical data remained unaware of the YiDiXie™ test results.

The study was approved by the Ethics Committee of Peking University Shenzhen Hospital and was conducted in accordance with the International Conference on Harmonization for “Good clinical practice guidelines” and the Declaration of Helsinki.

### Participants

This study included participants who had a positive CT plain for lung tumors. The two groups of subjects were enrolled independently, and each subject who satisfied the inclusion criteria was added one after the other.

Inpatients with “suspected malignancy” who had provided general informed consent for the donation of the remaining samples were initially included in the study. The study classified subjects into two groups based on their postoperative pathological diagnostic: those with a diagnosis of “malignant tumor” and those with a diagnosis of “benign disease”. The study omitted subjects whose pathology findings were not entirely clear. Some of samples in the malignant group were used in our prior works^11^.

This study did not include those who failed the serum sample quality test before the YiDiXie™ test. For information on enrollment and exclusion, please see the subject group’s prior article^11^.

### Sample collection, processing

The serum samples used in this study were obtained from serum left over after a normal medical consultation, which eliminated the need for additional blood sampling. To conduct the YiDiXie ™ test, 0.5 ml of serum was extracted from the individuals’ residual serum and stored at -80°C.

### The YiDiXie™ test

The YiDiXie ™ test utilizes the YiDiXie ™ all-cancer detection kit, an in vitro diagnostic kit developed and manufactured by Shenzhen KeRuiDa Health Technology Co. This test assesses the expression levels of several dozen miRNA biomarkers in serum to detect cancer in subjects. By integrating independent assays in a contemporaneous testing format and predefining suitable criteria for each miRNA biomarker, the YiDiXie ™ test maintains specificity and enhances sensitivity across a wide range of malignancies.

The YiDiXie ™ test comprises three distinct tests: YiDiXie ™ -Highly Sensitive (YiDiXie ™ -HS), YiDiXie ™ -Super Sensitive (YiDiXie ™ -SS), and YiDiXie ™ -Diagnosis (YiDiXie ™ -D). YiDiXie ™ -HS prioritizes specificity and sensitivity, while YiDiXie™ -SS significantly increases the number of miRNA tests to achieve high sensitivity across all clinical stages of various malignant tumor types. YiDiXie™ -Diagnosis (YiDiXie ™ -D) elevates the diagnostic threshold of a single miRNA test to ensure high specificity (low misdiagnosis rate) for all malignancy types.

The YiDiXie ™ test should be performed according to the instructions provided in the YiDiXie ™ all-cancer detection kit. Detailed procedures can be found in our prior works^11^. Following the completion of the test, the original results were analyzed by laboratory technicians from Shenzhen KeRuiDa Health Technology Co., Ltd, who determined the YiDiXie™ test outcomes as either “positive” or “negative”.

### Diagnosis of lung CT

The determination of “positive” or “negative” results is based on the diagnostic conclusion derived from the lung CT examination. A result is classified as “positive” if the diagnostic conclusion explicitly indicates malignancy. Conversely, if the diagnostic conclusion suggests a specific tumor type, leans towards benign interpretations, or includes phrases such as “probable malignant” or “further examination recommended”, the result is deemed “negative”.

### Extraction of clinical data

The hospitalized medical records or physical examination reports of the subjects were the source of the clinical, pathological, laboratory, and imaging data used in this investigation. Trained doctors assessed in accordance with the AJCC staging manual completed clinical staging. (7th or 8th edition)^12,13^.

### Statistical analyses

For the baseline qualities and demographic variables, descriptive statistics were given. The total number of individuals (n), mean, standard deviation (SD) or standard error (SE), median, first quartile (Q1), third quartile (Q3), minimum, and maximum values were used to describe continuous variables.

The number of individuals in each group and their proportion were used to express categorical variables. The 95% confidence intervals (CI) for the different indicators were determined using the Wilson scoring approach.

## RESULTS

### Participant disposition

This study ultimately included 1250 study subjects (malignant group, n = 1078; benign group, n = 172 cases). The demographic and clinical characteristics of the 1250 study subjects are listed in Table 1.

The two groups of study subjects were comparable in terms of demographic and clinical characteristics (Table 1). The mean (standard deviation) age was 52.7(13.15) years and 55.1%(689/1250) were female.

**Table 1.** Participant’s demographics and clinical manifestation.

|  | Malignant (n = 1078) | Benign (n = 172) | Total (N = 1250) |
| --- | --- | --- | --- |
| Age, years |  |  |  |
| Mean (SD) | 52.9 (13.27) | 52.0 (12.40) | 52.7 (13.15) |
| Median (Q1,Q3) | 53 (43, 64) | 54 (42, 62) | 53 (43, 63) |
| Min, max | 18, 88 | 20, 77 | 18, 88 |
| Age, group, n (%) |  |  |  |
| < 50 | 442 (41.0) | 75 (43.6) | 517 (41.4) |
| ≥ 50 | 636 (59.0) | 97 (56.4) | 733 (58.6) |
| < 65 | 833 (77.3) | 140 (81.4) | 973 (77.8) |
| ≥ 65 | 245 (22.7) | 32 (18.6) | 277 (22.2) |
| Sex, n (%) |  |  |  |
| Female | 619 (57.4) | 70 (40.7) | 689 (55.1) |
| Male | 459 (42.6) | 102 (59.3) | 561 (44.9) |
| Body mass index (kg/m<sup>2</sup>) |  |  |  |
| n | 1000 | 159 | 1159 |
| Mean (SD) | 23.2 (3.19) | 23.4 (3.25) | 23.2 (3.20) |
| Median (Q1,Q3) | 23.0 (21.1, 25.0) | 23.7 (21.1, 26.8) | 23.0 (21.0, 25.1) |
| Min, max | 14.8 45.2 | 16.5 32.5 | 14.8 45.2 |
| Body mass index category, n (%) |  |  |  |
| Underweight | 57 (5.3) | 10 (5.8) | 67 (5.4) |
| Benign | 564 (52.3) | 82 (47.7) | 646 (51.7) |
| Overweight | 311 (28.8) | 55 (32.0) | 366 (29.3) |
| Obese | 68 (6.3) | 12 (7.0) | 80 (6.4) |
| Missing | 78 (7.2) | 13 (7.6) | 91 (7.3) |
| AJCC clinical stage |  |  |  |
| Stage 0 | 306 (28.4) |  | 306 (28.4) |
| Stage I | 661 (61.3) |  | 661 (61.3) |
| Stage II | 69 (6.4) |  | 69 (6.4) |
| Stage III | 42 (3.9) |  | 42 (3.9) |

Q1,Q3, first quartile, third quartile; SD, standard deviation.

**Table 2.** Performance of YiDiXie ™-SS.

|  | Malignant | Benign | Total |
| --- | --- | --- | --- |
|  | 1078 | 172 | 1250 |
| Positive | 1051 | 57 | 1108 |
| Negative | 27 | 115 | 142 |

SEN, Sensitivity. SPE, Specificity. Two-sided 95% Wilson confidence intervals were calculated.

### Diagnostic performance of YiDiXie™-SS

As shown in Table 2, the sensitivity of YiDiXie™ SS was 97.5% (96.4% - 98.3%) and its specificity was 66.9% (59.5% - 73.5%). This means that YiDiXie™ SS has very high sensitivity and high specificity in lung tumors.

### Diagnostic performance of YiDiXie™-HS

As shown in Table 3, the sensitivity of YiDiXie™ -HS was 85.3% (83.0% - 87.2%) and its specificity was 83.1% (76.8% - 88.0%). This means that YiDiXie™-HS has high sensitivity and high specificity in lung tumors.

**Table 3.** Performance of YiDiXie^™^-HS.

|  | Malignant | Benign | Total |
| --- | --- | --- | --- |
|  | 1078 | 172 | 1250 |
| Positive | 919 | 29 | 948 |
| Negative | 159 | 143 | 302 |

SEN, Sensitivity. SPE, Specificity. Two-sided 95% Wilson confidence intervals were calculated.

### Diagnostic performance of YiDiXie™-D

As shown in Table 4, the sensitivity of YiDiXie™ -D was 73.9% (71.2% - 76.5%) and its specificity was 92.4% (87.5% - 95.5%). This means that YiDiXie™-D has high sensitivity and very high specificity in lung tumors.

**Table 4.**
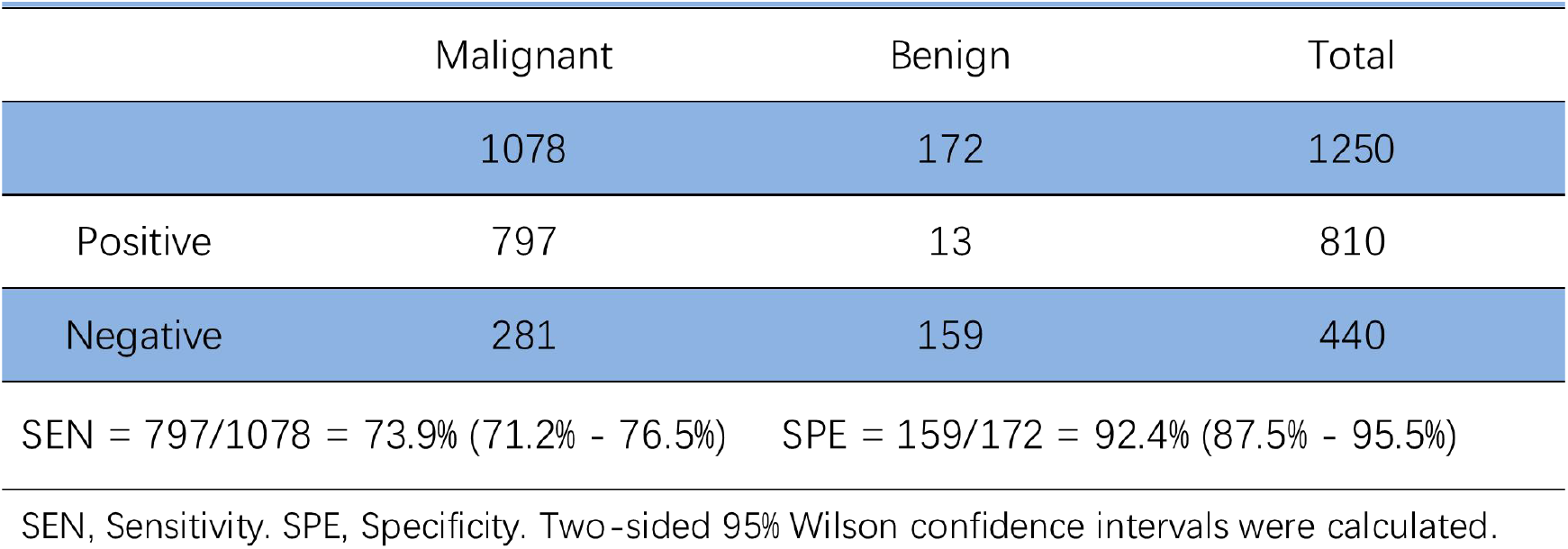
Performance of YiDiXie^™^-D.

### Diagnostic performance of lung CT

As shown in Table 5, the sensitivity of lung CT was 56.5%(95% CI: 53.5% - 59.4%) and its specificity was 72.7%(95% CI: 65.6% - 78.8%).

**Table 5.**
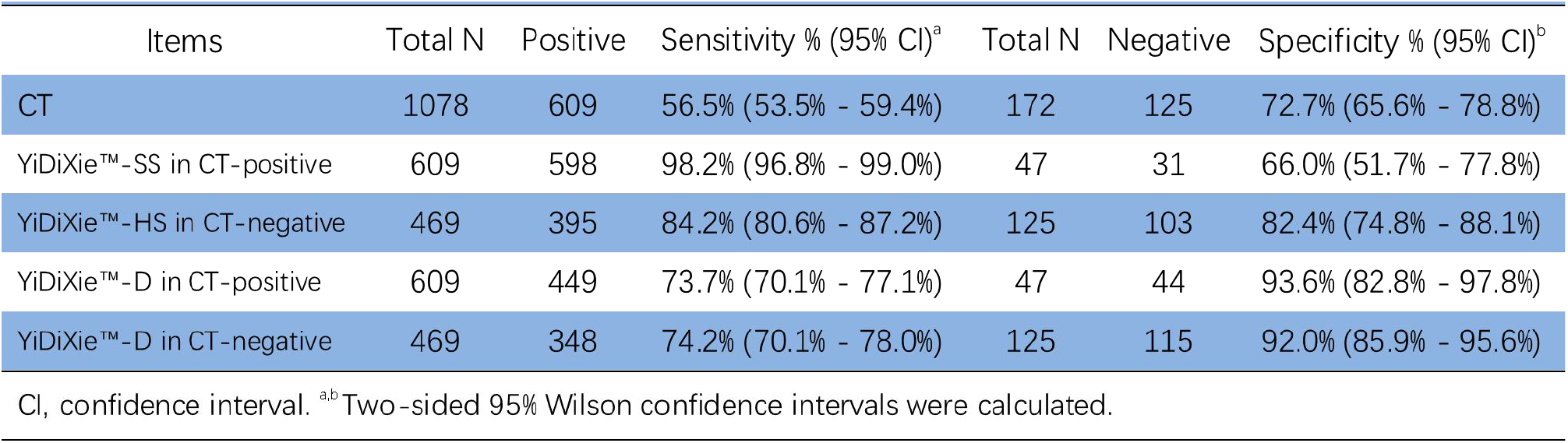
Performance of different Items.

### Diagnostic performance of in YiDiXie™-SS in lung CT-positive patients

In order to solve the challenge of high false-positive rate of lung CT, YiDiXie ™ -SS was applied to lung CT-positive patients.

As shown in Table 5, YiDiXie ™ -SS had a sensitivity of 98.2% (95% CI: 96.8% - 99.0%) and its specificity was 66.0% (95% CI: 51.7% - 77.8%) in patients with positive lung CT. This means that the application of YiDiXie ™ SS reduces the false-positive lung CT rate by 66.0% (95% CI: 51.7% - 77.8%) with essentially no increase in malignant tumor underdiagnosis.

### Diagnostic performance of YiDiXie™-HS in lung CT-negative patients

To address the challenge of high false-negative rate of lung CT, YiDiXie ™ -HS was applied to lung CT negative patients.

As shown in Table 5, YiDiXie ™ -HS had a sensitivity of 84.2% (95% CI: 80.6% - 87.2%) and its specificity was 82.4% (95% CI: 74.8% - 88.1%) in patients with negative lung CT. This means that the application of YiDiXie ™ -HS reduces the false negative rate of lung CT by 84.2% (95% CI: 80.6% - 87.2%).

### Diagnostic performance of YiDiXie™-D in lung CT-positive patients

False-positive consequences are significantly worse than false-negative consequences in some CT-positive patients, so YiDiXie™-D was applied to these patients to reduce their false-positive rates.

As shown in Table 5, YiDiXie ™ -D had a sensitivity of 73.7% (95% CI:70.1% - 77.1%) and its specificity was 93.6% (95% CI:82.8% - 97.8%) in patients with positive lung CT. This means that YiDiXie™-D reduces the false positive rate of lung CT by 93.6% (82.8% - 97.8%).

This means that YiDiXie ™ -SS reduces the false-positive lung CT rate by 95.7% (95% CI: 85.8% - 99.2%; 45/47).

### Diagnostic performance of YiDiXie™-D in lung CT-negative patients

Certain CT-negative patients had significantly worse false-positive than false-negative consequences, so the more specific YiDiXie ™ -D was applied to such patients.

As shown in Table 5, YiDiXie ™ -D has a sensitivity of 74.2% (95% CI:70.1% - 78.0%) and its specificity is 92.0% (95% CI:85.9% - 95.6%) in patients with negative lung CT. This means that YiDiXie™-D reduces the false-negative rate of lung CT by 74.2% (95% CI:70.1% - 78.0%) while maintaining high specificity.

## DISCUSSION

### Clinical significance of YiDiXie™-SS in lung CT-positive patients

The YiDiXie ™ test comprises three tests with distinct characteristics: “YiDiXie™-Highly Sensitive” (YiDiXie™-HS), “YiDiXie™-Super Sensitive” (YiDiXie ™ -SS) and “YiDiXie ™ -Diagnosis” (YiDiXie ™ -D). Among these, YiDiXie™-HS offers a balance of high sensitivity and specificity, while YiDiXie ™ -SS prioritizes high sensitivity for all malignant tumor types at the expense of slightly lower specificity.

Given the clinical importance of both sensitivity and specificity in patients with positive CT scans for lung tumors, the selection of further diagnostic methods involves a trade-off between the risk of missing malignant tumors and the risk of misdiagnosing benign tumors. Typically, benign lung tumors often will likely lead to unnecessary surgery when misdiagnosed as malignant, but it will not affect the patient’s prognosis, and its treatment is much less expensive than that of advanced cancers. Furthermore, its positive predictive value is higher in lung CT-positive patients. The false-negative rate can be more harmful even if it is comparable to the false-positive rate. Consequently, YiDiXie ™ -SS, with its very high sensitivity but slightly lower specificity, was chosen to mitigate lung CT false-positive rate.

As shown in Table 5, YiDiXie ™ -SS had a sensitivity of 98.2% (95% CI: 96.8% - 99.0%) and its specificity was 66.0% (95% CI: 51.7% - 77.8%) in lung CT-positive patients. The above results suggest that YiDiXie™-SS reduces the false-positive rate of lung CT by 66.0% (95% CI: 51.7% - 77.8%) while maintaining a sensitivity close to 100%.

These results imply that YiDiXie ™ -SS substantially reduces the probability of incorrectly surgery for benign lung disease without essentially increasing the number of missed diagnoses of malignant tumors. In other words, YiDiXie ™ -SS substantially reduces the mental suffering, expensive examination and surgical costs, surgical injuries, and other adverse consequences for patients with false-positive lung CT scans, without essentially increasing the number of cases of delayed treatment of malignant tumors. Therefore, YiDiXie™-SS well meets the clinical needs and has important clinical significance and wide application prospects.

### Clinical significance of YiDiXie™-HS in lung CT-negative patients

For patients with negative lung CT scans, both sensitivity and specificity of further diagnostic methods are crucial. Balancing this contradiction between sensitivity and specificity essentially involves weighing the “risk of missing malignant tumors” against the “risk of misdiagnosing benign diseases”. A higher false-negative rate implies more missed malignant tumors, leading to delayed treatment, disease progression, and potentially advanced-stage malignancies. Patients may then suffer from poorer prognosis, shorter survival, decreased quality of life, and higher treatment costs. Generally, when benign lung tumors are misdiagnosed as malignant, they are typically managed with surgery without affecting patient prognosis, with treatment costs significantly lower than those for advanced cancer. Therefore, for patients with negative lung CT scans, the “risk of missing malignant tumors” outweighs the “risk of misdiagnosing benign diseases”. Hence, the choice of YiDiXie™-HS with high sensitivity and specificity is selected to reduce the false-negative rate of lung CT scans for lung tumors.

As shown in Table 5, the sensitivity of YiDiXie™ -HS in lung CT-negative patients was 84.2% (95% CI: 80.6% - 87.2%) and its specificity was 82.4% (95% CI: 74.8% - 88.1%). The above results suggest that the application of YiDiXie ™ -HS reduced the false-negative rate of lung CT by 84.2% (95% CI: 80.6% - 87.2%).

These findings imply that YiDiXie ™ -HS significantly reduces the probability of missed malignant tumors in patients with negative lung CT scans. In other words, YiDiXie ™ -HS substantially decreases the negative consequences such as poorer prognosis, higher treatment costs, decreased quality of life, and shorter survival in patients with false-negative lung CT scans for lung Therefore, YiDiXie ™ -HS effectively meets clinical needs, offering significant clinical significance and promising application prospects.

### Clinical significance of YiDiXie™-D

Lung tumors considered malignant are usually treated surgically. However, there are some conditions that require extra caution in choosing whether to operate or not, hence further diagnosis, e.g., smaller tumors, tumors that require lobectomy, patients with significant pulmonary insufficiency, patients in poor general condition, etc.

In patients with lung tumors, both the sensitivity and specificity of further diagnostic approaches are important. Weighing the contradiction between sensitivity and specificity is essentially weighing the contradiction between the “harm of malignant tumor underdiagnosis” and the “harm of benign tumor misdiagnosis”. Because smaller tumors have a lower risk of tumor progression and distant metastasis, the “risk of malignant tumor underdiagnosis” is much lower than the “risk of benign tumor misdiagnosis”. For tumors requiring lobectomy, the risk of misdiagnosis of benign tumors is much higher than the risk of misdiagnosis of malignant tumors because the affected lobe of the lung needs to be removed. For patients with obvious pulmonary insufficiency, the risk of misdiagnosis of benign tumors is much higher than that of malignant tumors because of the likelihood of postoperative dyspnea. For patients with poor general conditions, the “risk of misdiagnosis of benign tumors” is much higher than the “risk of malignant tumors” because the perioperative risk is much higher than the general condition. Therefore, For these patients, YiDiXie™-D, which is highly specific but slightly less sensitive, was chosen to reduce the false-positive rate of lung CT or to significantly reduce its false-negative rate while maintaining high specificity.

As shown in Table 5, the sensitivity of YiDiXie™ -D in lung CT-positive patients was 73.7% (70.1% - 77.1%), and its specificity was 93.6% (82.8% - 97.8%). YiDiXie ™ -D in lung CT-negative patients had a sensitivity of 74.2% (70.1% - 78.0%) and a specificity of 92.0% (85.9% - 95.6%). - 95.6%). These results indicate that YiDiXie™-D reduces the false-positive lung CT rate by 93.6% (82.8% - 97.8%) or reduces the false-negative lung CT rate by 74.2% (70.1% - 78.0%) while maintaining a high specificity.

These results imply that YiDiXie ™ -D significantly reduces the likelihood of incorrect surgery in these patients who require extra caution. In other words, YiDiXie ™ -D significantly reduces the risk of adverse outcomes such as surgical trauma, organ resection, pulmonary insufficiency, ventilator maintenance, and even death and other serious perioperative complications in these patients. Therefore, YiDiXie ™ -D well meets the clinical needs and has important clinical significance and wide application prospects.

### YiDiXie™ test has the potential to solve two challenges of lung tumor

First, the three products of the YiDiXie™ test are clinically important in lung tumors. As mentioned earlier, the YiDiXie ™-SS, YiDiXie ™ -HS and YiDiXie™-D have significant diagnostic value in patients with positive or negative enhanced CT, respectively.

Second, the three products of YiDiXie ™ test can greatly relieve the work pressure of clinicians tumors. and promote timely diagnosis and timely treatment of malignant tumor cases that would otherwise be delayed. On the one hand, YiDiXie™-SS also offers significant relief from non-essential work for surgeons. Positive enhanced CT lung tumors are often treated surgically. Whether these surgeries can be completed in a timely manner is directly dependent on the number of surgeons.

Appointments are booked for months or even more than a year in many parts of the world. A delay in the treatment of these malignant cases is inevitable, and thus malignant progression or even distant metastasis is not uncommon in patients with lung tumors awaiting surgery. As shown in Table 5, YiDiXie™-SS reduced the lung CT false-positive rate by 66.0% (95% CI: 51.7% - 77.8%) with essentially no increase in lung cancer leakage. Thus, YiDiXie ™ -SS can significantly reduce the non-essential workload of surgeons, facilitating timely diagnosis and treatment of lung tumors or other diseases that would otherwise be delayed.

On the other hand, YiDiXie™-HS and YiDiXie™ -D can greatly reduce clinicians’ work pressure. When the diagnosis is difficult on lung CT, the patient usually requires an enhanced MRI or a puncture biopsy. The timely completion of these enhanced MRIs or puncture biopsies is directly dependent on the number of imaging physicians available. Appointments are available for months or even more than a year in many parts of the world. It is also not uncommon for patients with lung tumors waiting for an enhanced MRI exam or puncture biopsy to experience malignant progression or even distant metastases. YiDiXie ™ -HS and YiDiXie ™-D can replace these enhanced MRI examinations or puncture biopsies, greatly easing clinicians’ workload and facilitating timely diagnosis and treatment of other tumors that would otherwise be delayed.

Final, the YiDiXie ™ test can achieve “timely diagnosis” of lung tumors. On the one hand, the YiDiXie™ test requires only a tiny amount of blood and allows patients to complete the diagnostic process non-invasively without having to leave their homes. Only 20 microliters of serum are required to complete a YiDiXie ™ test, which is equivalent to approximately one drop of whole blood (one drop of whole blood is approximately 50 microliters, which yields 20-25 microliters of serum)^11^. Taking into account the pre-test sample quality assessment test and 2-3 repetitions, 0.2 ml of whole blood is sufficient for the YiDiXie™ test^11^. The 0.2 ml of finger blood can be collected at home by the average patient using a finger blood collection needle, eliminating the need for venous blood collection by medical personnel, and allowing patients to complete the diagnostic process non-invasively without having to leave their homes^11^.

On the other hand, the diagnostic capacity of the YiDiXie ™ test is virtually unlimited. Figure 1 shows the basic flowchart of the YiDiXie ™ test, which shows that the YiDiXie™ test not only does not require a doctor or medical equipment, but also does not require medical personnel to collect blood.

**Figure 1.**
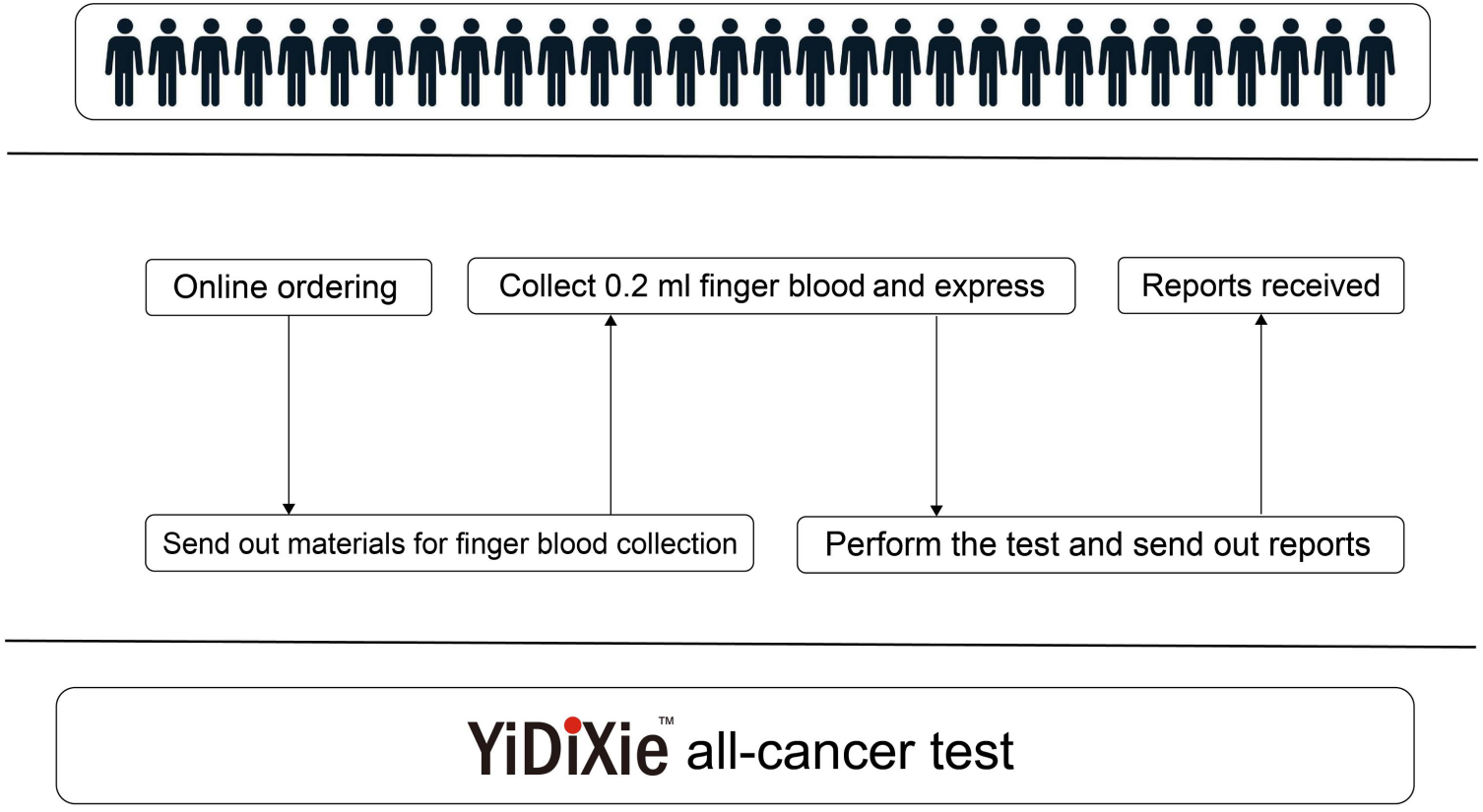
Basic flowchart of the YiDiXie™ test.

As a result, the YiDiXie ™ test is completely independent of the number of medical personnel and facilities, and has a virtually unlimited test capacity. In this way, the YiDiXie ™ test enables “just-in-time” diagnosis of lung tumors without patients having to wait anxiously for an appointment.

In short, the YiDiXie ™ test has significant diagnostic value in lung cancer, and is expected to solve the problems of “ high false-positive rate of lung CT” and “high false-negative rate of lung CT”.

### Limitations of the study

Firstly, the number of cases in this study was small, and future clinical studies with larger sample sizes are needed for further evaluation.

Secondly, this study was a malignant tumor case-benign tumor control study in hospitalized patients, and future cohort studies in the natural population of lung tumors are needed for further Finally, this study was a single-center study, which may have led to some degree of bias in the results of this study. Future multicenter studies are needed for further evaluation.

## CONCLUSION

YiDiXie ™ -SS provides extremely high sensitivity and high specificity in lung tumors. YiDiXie ™ -HS provides high sensitivity and high specificity in lung tumors. yiDiXie ™ -D provides high sensitivity and very high specificity in lung tumors. YiDiXie™-SS significantly reduces the rate of lung CT false positives with essentially no increase in delayed treatment of malignant tumors. YiDiXie ™ -HS significantly reduces the false-negative rate of lung CT. YiDiXie ™ -D can significantly reduce the false-positive rate of lung CT or significantly reduce the false-negative rate while maintaining high specificity. the YiDiXie™ test has important diagnostic value in lung cancer and is expected to solve the problems of “ too high false-positive rate “ and “ too high false-negative rate” of lung CT.

## Data Availability

All data produced in the present study are contained in the manuscript.

